# Coding of Obesity-related Mortality Impacts Estimates of Obesity on U.S. Life Expectancy

**DOI:** 10.1101/2022.05.16.22275140

**Authors:** Andrea M. Tilstra, José Manuel Aburto, Iliya Gutin, Jennifer Beam Dowd

## Abstract

**Background:** High levels of obesity remain an important population health problem in the U.S. and a possible contributor to stalling life expectancy. However, reliable estimates of the contribution of obesity to mortality in the U.S. are lacking, because of inconsistent coding of obesity-related causes of death.

**Methods:** We compare five International Classification of Diseases version 10 (ICD-10) coding schemes for obesity-related mortality used in the literature and examine how the magnitude of obesity-related mortality burdens varies across different schemes. We use U.S. multiple cause of death data and population estimates for the Black, white, and Latino population in the years 2010, 2015, and 2020. In sex- and race/ethnic-stratified analyses, we estimate the potential years of life expectancy gained if obesity-related mortality had not occurred as measured by each coding scheme.

**Results:** We estimate that obesity-related mortality contributes to up to 78 months (6.5 years) of lost U.S. life expectancy, though estimates range from as low as 0 months, with a median contribution across ICD-10 coding schemes of about 20 months (1.7 years). Despite substantial variation across coding schemes, obesity-related mortality consistently contributes more to life expectancy deficits for Black Americans compared to white and Latino Americans. Across all ICD-10 coding schemes, the age pattern of obesity follows a J-shaped curve, suggesting exponential increases in obesity-related mortality after age 25.

**Conclusions:** The estimation of the burden of obesity-related mortality on life expectancy in the United States varies widely depending on the causes of death used in analyses. This inconsistency may obscure our understanding of the contribution of obesity-related mortality to trends in life expectancy. We propose a standardization of the coding of obesity-related mortality for future studies and outline which causes should be included.

## Introduction

In 2018, an estimated 42.5% of the United States population had obesity (body mass index (BMI) ≥30), an increase from 30.5% in 2000. Over the same period, rates of severe obesity (BMI≥ 40) nearly doubled to 9.2% (1). There are pronounced race/ethnic differences in American obesity rates, as evidenced by the 57% obesity rate among non-Hispanic Black women compared to 40% among non-Hispanic white women (1). While the obesity epidemic defies easy explanation, one hypothesis is that an increasingly obesogenic environment over past decades (e.g., increased accessibility of calorically-dense food and more sedentary lifestyles) has made maintaining a healthy body weight more challenging, in ways that are socially patterned (2). Consequently, the obesity epidemic may be contributing to alarming U.S. mortality and life expectancy trends (3–5), as well as racial disparities in these trends (6,7). Research also indicates that these trends may continue to worsen as younger cohorts are exposed to more time in the U.S. obesogenic environment (3,8).

Despite the increased prevalence of obesity in the United States and widespread recognition of its consequences for population health, we lack consensus on how to track and report its impact on key population health metrics such as mortality rates and life expectancy estimates. As argued in this paper, a key obstacle in obtaining such estimates is the absence of a standardized approach for measuring obesity-related mortality based on reporting standards in existing vital statistics data.

Currently, statistical approaches for estimating obesity-related mortality generally fall into one of two categories based on the data source used for analysis. One approach uses obesity prevalence and all-cause mortality to estimate obesity-attributable mortality fractions (OAMF) (9). To estimate this, researchers use nationally-representative survey data (e.g., National Health Interview Survey, National Health and Nutrition Examination Survey) to calculate the prevalence of obesity and the relative risk of premature mortality associated with obesity as compared to a lower weight status. The two are then combined to provide a counterfactual proportion of deaths attributable to obesity that could have otherwise been avoided if all adults were assigned to a lower weight status (10). There are different techniques to estimate OAMF, including the partially adjusted method and the weighted sum method (11,12). Yet, the OAMF approach is often limited in estimating an association between obesity and/or BMI at a given point in time and mortality many years later, as there are many possible sources of confounding that introduce substantial uncertainty in the meaning of a single measure of obesity (13–15).

A second, more recent approach – which is the subject of this analysis – uses underlying and multiple cause of death mortality data to estimate obesity-related mortality rates (8,16–18). Because of the very strong and likely causal association between obesity and a broad variety of cardiometabolic conditions, many different causes of death are often categorized as obesity-related (9,19–23). Though categorizations from multiple cause of death data cannot definitively capture obesity-related mortality, they may be a better proxy for such deaths and provide more “direct individual-level evidence” given that they come from death certificates rather than OAMFs from survey data (16). These data are also valuable in highlighting the many co-morbid conditions that contribute to adult mortality, as the information provided by a single, underlying cause may be insufficient (24–26). While the demographic analysis techniques used with population-level cause-specific mortality data are less varied across studies (e.g., estimation of age-standardized mortality rates), there is a clear lack of consistency in the classification of obesity- or metabolic-related causes of death. The relatively standardized statistical approaches with population-level mortality data warrant a standardized coding scheme for consistent and comparable estimates of obesity-related mortality across studies, time, and subpopulations of interest.

The International Classification of Disease (ICD) lists obesity as an independent cause of death (ICD10: E65-E67), but researchers acknowledge that this coding is used too infrequently and underestimates the true mortality burden of obesity, as there are many other causes of death which are likely representative of obesity-related mortality, including hypertension and diabetes (27–29). Thus, studies using multiple cause of death data often include additional ICD codes in their obesity coding schemes. However, we contend that the inconsistent use of ICD codes across studies is a key limitation of this work as it leads to vastly different estimates of the contribution of obesity to U.S. mortality.

In this paper, we test how different obesity-related ICD coding schemes equate to variation in the estimated of years of life expectancy in the U.S. lost due to obesity. We also show how these patterns differ by sex and race/ethnicity, given large disparities in obesity rates within the U.S. population. Finally, we identify critical flaws in commonly used obesity-related ICD coding schemes and propose best practices for standardizing. In turn, our systematic comparison of these different coding practices – both over time and by sex and race/ethnicity – helps demonstrate the considerable variation in obesity’s impact over past decades and across groups. Ultimately, we argue that greater standardization is vital for improving our knowledge of population health trends in obesity-related mortality and its impact on life expectancy.

## Data & Methods

### Data

Data come from the publicly available United States mortality multiple cause of death files provided by the Vital Statistics division of the National Center for Health Statistics (numerator) and yearly population estimates from the Surveillance, Epidemiology, and End Results Program (denominator) (30). We restrict data to U.S. residents in the years 2010, 2015, and 2020, and to non-Hispanic white (white), non-Hispanic Black (Black), and Hispanic/Latino (Latino) populations. Other race/ethnic groups were excluded due to issues with model instability from small data counts. All analyses are sex- and race/ethnic-stratified.

### Coding Schemas

We identify five coding schemes for obesity-related mortality. Three come from recent publications: Masters et al. (8) (henceforth: Masters), Adair and Lopez (16) (henceforth: Adair), and Acosta et al. (18) (henceforth: Acosta). The fourth comes from the Global Burden of Disease (GBD) 2013 estimates of mortality attributable to high BMI (17), and the fifth coding scheme uses only the ICD codes for obesity (E65-E67) as the underlying cause of mortality (UCOD). Appendix A includes the ICD-10 codes for each coding scheme. Several differences across coding schemes are worth noting. First, GBD and Acosta are the only two to include neoplasms. Acosta restricts to “obesity-related” neoplasms, but GBD codes are much more expansive (e.g., leukemia). Second, Acosta and UCOD do not include any hypertension or heart disease causes, while the other three coding schemes do.

### Other Causes of Death

To show the relative contribution of obesity-related deaths compared to other major causes of death shaping recent life expectancy trends in the United States, we code and analyze five major causes of mortality and one residual category. These are despair-related mortality (suicides, alcohol- and drug-related) (8), cancer deaths, deaths from infectious and parasitic diseases, deaths from respiratory diseases, and accidental deaths (excluding those related to despair). The ICD-10 codes used in each category are listed in Appendix B. All categories are mutually exclusive, such that if a cause of death was listed in an obesity scheme and in another category, its inclusion in the obesity category was prioritized. For example, the obesity-related neoplasms that were included in Acosta scheme were not included in cancer mortality for analyses of the Acosta scheme.

### Methods

Lifetables were calculated by sex and racial/ethnic groups following standard demographic techniques (31). From these, life expectancy at birth was used as the main indicator of our analysis. Life expectancy is the average number of years a cohort of newborns is expected to live if they were to experience the mortality rates present in a given year throughout their lives. It is not a forecast or projection of any individual’s lifespan, but it accurately reflects mortality levels of a population in a given year. Because life expectancy is not affected by population size or age-structures, it is a comparable indicator across populations and over time.

To calculate the burden of obesity-related mortality on life expectancy in a comparable way across populations and schemes, we used cause-deleted life tables and calculated the potential years of life expectancy that would be gained if obesity-related deaths were avoided (31). This methodology has been used previously in similar studies and is similar to decomposition techniques used in demographic research (32).

## Results

Figures 1 and 2 show the distribution of mortality across causes of death for the United States in 2020 for females (Figure 1) and males (Figure 2). While the residual category consistently comprises the most deaths, despair-related mortality and accidents stand out as the key contributors of mortality at middle-ages. Despair-related deaths are especially pronounced among middle-aged U.S. men of all races/ethnicities. These figures show that the prevalence of obesity-related mortality increases with age. Under the UCOD scheme there are very few obesity-related deaths (less than 0.6%), across both sexes and all three race/ethnic categories. While under the GBD coding scheme it comprises a large proportion of deaths, from 28.5% for Latino males to 37.0% among Black females. Across all schemes, there is a higher proportion of obesity-related deaths for Black males and females than for white and Latino males and females, consistent with racial and ethnic disparities in obesity prevalence.

**Figure 1.**
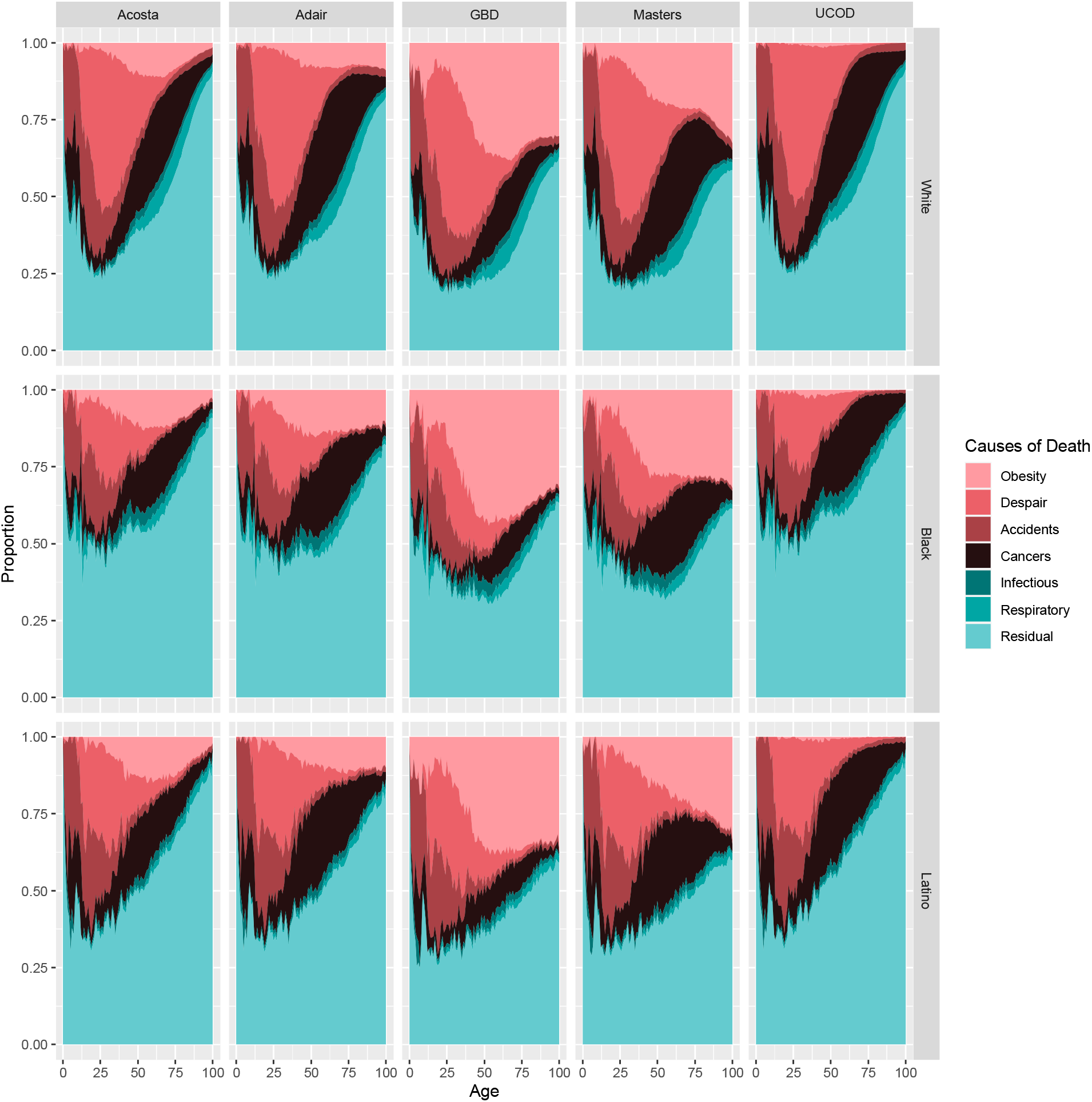
Cause of Death Structure over Age, Coding Scheme, and Race/Ethnicity, Females 2020. Notes: Data come from the NVSS mortality data and the SEER population estimates.

**Figure 2.**
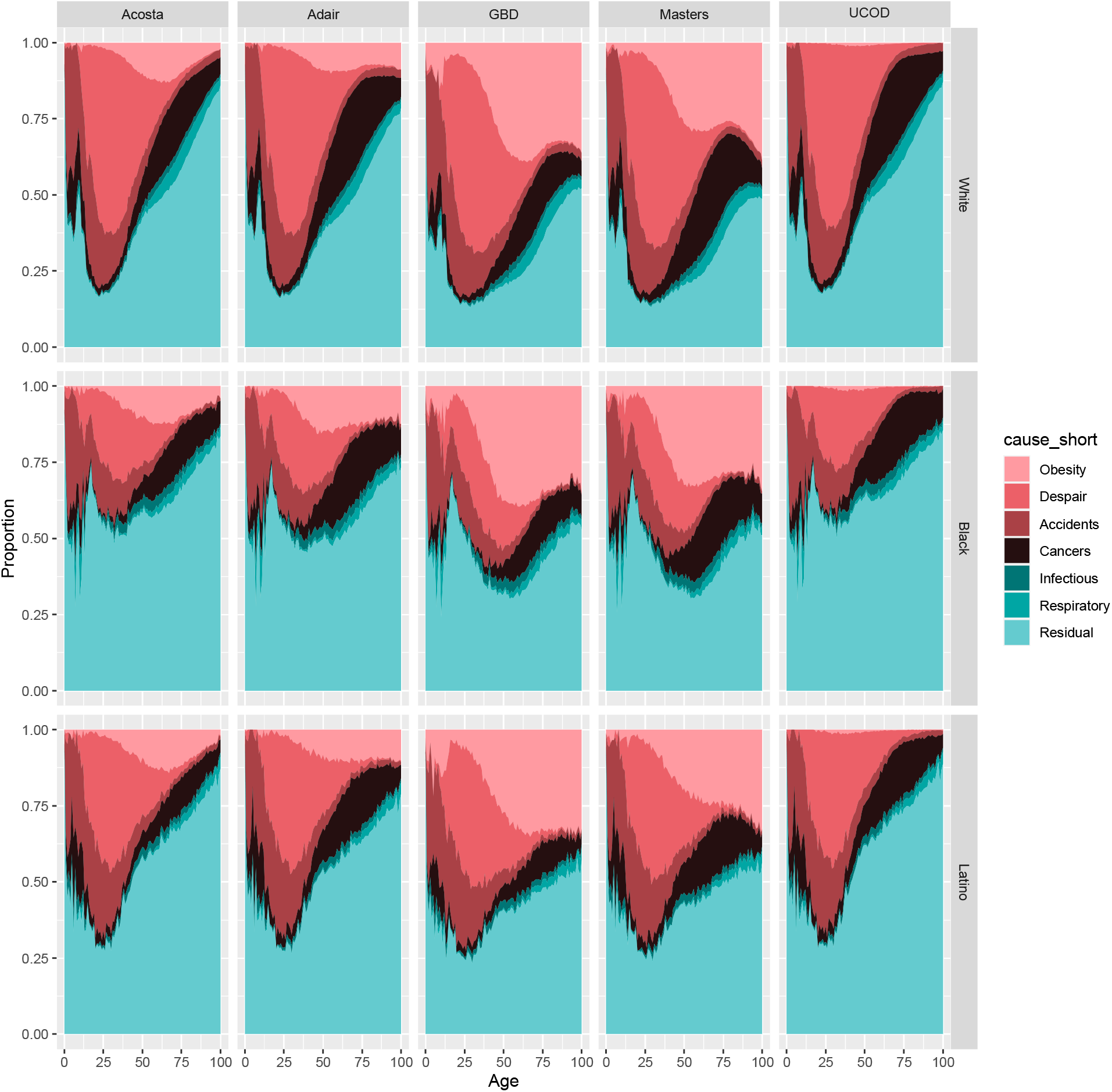
Cause of Death Structure over Age, Coding Scheme, and Race/Ethnicity, Males 2020. Notes: Data come from the NVSS mortality data and the SEER population estimates.

The age pattern of obesity-related mortality, shown in Figure 3, is relatively consistent across coding schemes. It is also consistent over time (as shown in Appendix C). The J-shaped curve suggests that obesity-related mortality increases exponentially with age after around age 25, showing that even at young ages (0-20), people are dying from causes deemed to be obesity-related, albeit at much smaller rates. The exception is, again, the UCOD scheme which shows little to no obesity-related deaths at young ages. This scheme also shows a stable distribution across ages, suggesting that the likelihood of a death being coded with obesity as the underlying cause does not fluctuate much across ages. At older ages, the Acosta scheme deviates from other trajectories, which may be indicative that it more accurately captures frailty or selection.

**Figure 3.**
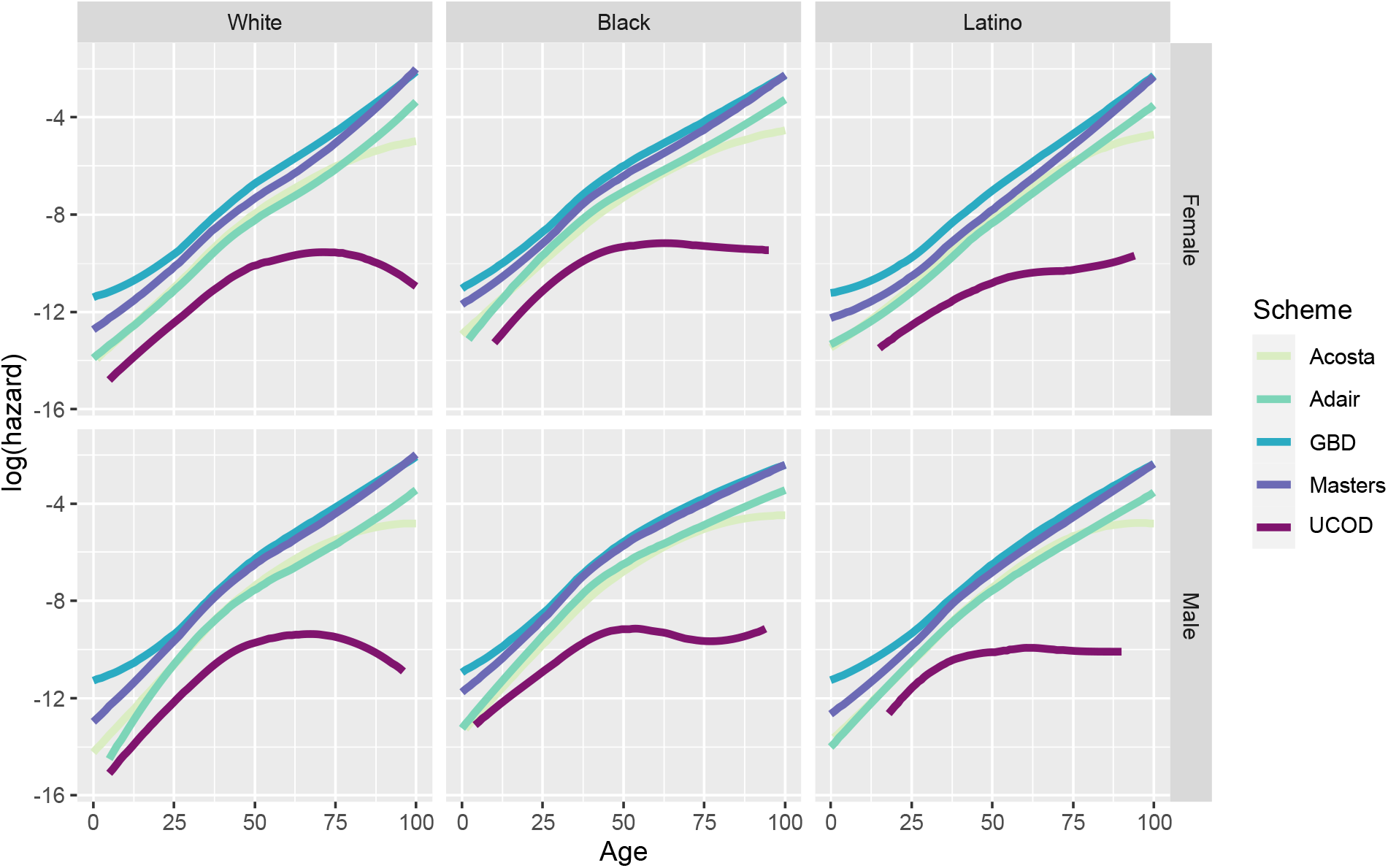
Age Pattern of Obesity-related Mortality across Schemas, 2020. Notes: Data come from the NVSS mortality data and the SEER population estimates.

Table 1 provides estimates of life expectancy at birth (e_0_) for females and males in 2010, 2015, and 2020. Consistent with estimates provided by the U.S. Vital Statistics (33), Black males have the lowest life expectancy (67.79 years in 2020) and male and female Latinos have life expectancy that is consistently higher than their white and Black counterparts. Additionally, estimates of life expectancy dropped between 2015 and 2020 for all sex and race/ethnic groups, except white women.

**Table 1.** Life Expectancy at Birth (e_0_) by Race/Ethnicity and Sex: 2010, 2015, and 2020

|  | Female |  |  | Male |  |  |
| --- | --- | --- | --- | --- | --- | --- |
|  | 2010 | 2015 | 2020 | 2010 | 2015 | 2020 |
| **White** | 81.10 | 81.03 | 80.21 | 76.36 | 76.33 | 74.93 |
| **Black** | 77.64 | 78.11 | 75.52 | 71.40 | 71.72 | 67.79 |
| **Latino** | 84.57 | 84.81 | 82.34 | 79.69 | 80.04 | 75.62 |

Notes: Data come from the NVSS mortality data and the SEER population estimates.

Figure 4 shows race/ethnic and sex differences in the contribution of obesity-related mortality to life expectancy estimates in 2010, 2015, and 2020. The contribution of obesity-related causes to estimates of life expectancy vary widely across coding schemes from 0 months (UCOD) to 80 months (GBD). Consistent across schemes is that obesity-related mortality contributed more to losses in Black American life expectancy than white and Latino life expectancy. For instance, in 2020 the GBD coding scheme showed that obesity contributed 78 months to lost Black female life expectancy, and 64 and 53 months to white and Latina females, respectively. In parallel, eliminating obesity-related mortality under the GBD coding scheme would result in the largest improvements in U.S. life expectancy. Yet, contributions of obesity-related mortality to life expectancy losses for all schemes (except UCOD) exceed 10 months. Of note, contributions are similar for 2010 and 2015, but lower in 2020; this may be a result of an increase in deaths from COVID-19 during 2020.

**Figure 4.**
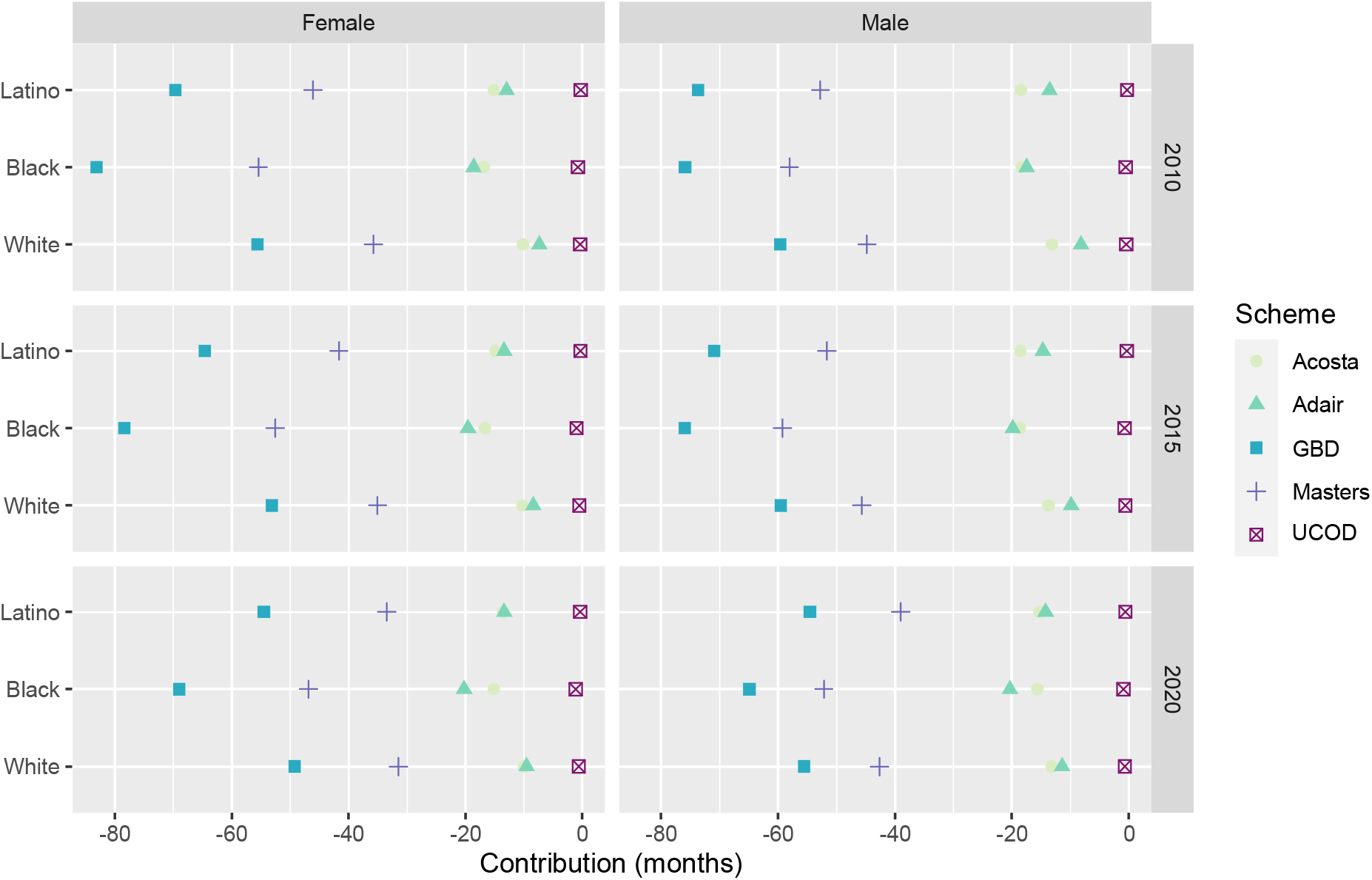
Contribution of Obesity to Life Expectancy losses by Coding Schema: 2010, 2015, and 2020. Notes: Data come from the NVSS mortality data and the SEER population estimates.

## Discussion

The high and rising rates of obesity in the U.S. suggest that the effect of obesity on U.S. life expectancy will continue to be significant. Understanding the contribution of obesity to stalls in U.S. life expectancy and its deterioration relative to peer countries is an important area of research (10,12,16,18). Thus, the measurement of obesity-related mortality is both substantively and theoretically important, but approaches have not been standardized up to now. Our results suggest several key takeaways for moving forward.

First, we find that the elimination of obesity-related mortality would increase life expectancy for all Americans, regardless of the coding scheme used. While the effect size differs depending on the scheme, we show that if these causes were eliminated, life expectancy could improve by as much as 80 months, or 6.6 years. Second, we find that the elimination of obesity-related mortality would increase life expectancy more for Black Americans than for other racial/ethnic groups in the U.S. The contribution of obesity-related mortality is also greater at younger ages for Black Americans, compared to their white and Latino counterparts. This is likely a result of complex social determinants of body weight, which particularly disadvantage the Black population in early life and throughout the life course (34–37).

This study has important implications for the measurement of obesity-related mortality. First, we recommend that UCOD, or obesity as the single underlying cause of death, should not be used for estimating the contribution of obesity to mortality in a population. It is an overly narrow definition of obesity-related deaths, which necessitates that obesity is listed as not only a cause of death on the death certificate, but as the underlying cause. Unfortunately, this strict criterion is potentially subject to the biases/assumptions of the person coding the cause of death (27,29), leading to the identification of less than a fifth of all deaths where obesity is implicated (24). For instance, a cross-national comparison of obesity-related mortality suggests that physicians’ propensity for reporting obesity as an underlying cause on death certificates might vary due to their “sensitiv[ity] to abnormally high BMIs,” in reference to national differences in the prevalence of obesity and thus expectations about the *average* BMI and body size (24). Within the context of the United States, reporting practices may be similarly influenced by sex and racial/ethnic differences surrounding body size norms and health (38–40), as well as sex and racial/ethnic biases in how medical practitioners view their patients and health (41–43). These factors may in turn influence perceptions about obesity’s role for individuals’ health (44,45), and thus its contribution to their death. Thus, it is evident that existing coding practices for obesity as an underlying cause are not only too limited in identifying obesity-related comorbidity, but also likely more susceptible to the subjective preferences and beliefs of the individual evaluating the reasons underlying death.

Conversely, the GBD coding scheme is likely too broad. It includes several types of cancers (e.g., leukemia and ovarian cancer) which, while linked with obesity, are likely not a direct result of obesity. We suggest that future measurements of obesity-related mortality consider adopting the Adair coding scheme. This scheme, along with the Acosta scheme, provide moderate yet plausible estimates for the contributions of obesity-related mortality for life expectancy. Yet unlike Acosta and Masters schemes, it includes causes of death that are more likely to be a direct result of obesity: obesity (included in Adair, Acosta, Masters, UCOD), diabetes (Adair, Acosta, GBD, Masters), hypertension (Adair, Masters), chronic kidney disease (Adair, GBD), and lipidemia-related causes (Adair, GBD). The exact ICD-10 codes used the Adair scheme, and all other schemes, are provided in Appendix A. It is inevitable that there is some margin of error in how well each of these causes of deaths can be directly attributable to obesity, but past studies clearly demonstrate where and when this association is stronger or weaker (9,17,21,46). Future research might consider developing a relative risk threshold for the association between obesity and specific causes of death, to use direct epidemiologic evidence to determine which causes to include. For now, the codes used by Adair and Lopez strike the best balance of including conditions most strongly associated with obesity (16).

Second, the age pattern of mortality across coding schemes emphasizes the importance of considering when in the life course obesity can plausibly be considered a contributing cause of death. Obesity has relatively immediate health consequences, mostly reflected in its association with elevated biomarkers for poor cardiovascular and metabolic health (47–49), as well as its broad pro-inflammatory effect (50,51), which is implicated in numerous chronic conditions. However, the health consequences that might increase the risk of mortality are typically cumulative in nature – especially with respect to chronic cardiometabolic disease – and thus unlikely to be more directly implicated until later in life, when adults have lived with one or more chronic diseases for an extended period of time (52–54). Moreover, some proportion of obesity-related deaths are likely misclassified when using multiple cause of death data. This misclassification bias may be more salient at younger ages where even causes of death that are very strongly linked to obesity (e.g., diabetes, heart disease) have a more complex etiology. Thus, it is unlikely that young individuals are dying directly of obesity-related mortality. To account for this, we suggest that researchers consider restricting to ages 25 and older when estimating obesity-related mortality research. This is particularly important for research estimating the consequences of obesity-related mortality on life expectancy which, unless specified otherwise, considers all ages in the estimation.

### Limitations

As noted earlier, the only currently-available ICD coding options that *specifically* implicates obesity as the underlying cause of death are the E66-67 codes, which likely heavily undercount obesity-related deaths – as demonstrated in this analysis and suggested in earlier work (16,27– 29). Thus, it is inevitable that at least some proportion of the deaths in each scheme are not due to obesity, but instead a function other lifestyle and health factors, such as smoking (17). It is difficult to say whether the estimates presented by different coding schemes are liberal or conservative, as any “overestimation” noted above may be balanced out by “underestimation” in obesity’s association with numerous other causes of death not included in the schemes (e.g., infectious disease [(24)]).

It is worth reiterating that the consequences of obesity for mortality can be estimated with both population-level data and demographic techniques, as is shown here, or with obesity-attributable mortality functions (OAMF). OAMFs, while useful because they estimate the direct relationship between obesity and mortality, are flawed because they estimate population-level obesity-related mortality using only a sample of the population, whose obesity was estimated at a time point preceding their mortality. Additionally, there is some evidence that the BMI cutoffs for obesity are perhaps not the same across all sex and race/ethnic groups, with a weaker association between high BMI and mortality for non-Hispanic Black and Hispanic men (55–58). In turn, there may be nonnegligible variation in how accurately multiple cause of death coding schemes capture obesity-related mortality (i.e., overestimation or underestimation) across different subpopulations. However, studies are largely equivocal in reaching consensus on the (in)appropriateness of traditional BMI categories across the population and how obesity is defined, so it is unclear how exactly this might affect the estimates presented in our analysis.

## Conclusions

This paper fills an important gap in obesity and mortality research by demonstrating the consequences of different coding schemes for estimates of life years lost due to obesity-related mortality. The approach of examining causes of death where obesity is not explicitly the underlying cause mirrors efforts to estimate the “true” burden of mortality associated with other risk factors for disease, like smoking (59). Indeed, we argue that researchers should be aware of the broad range of plausible estimates for obesity-related mortality and resulting deficits in life expectancy, as a single estimate is likely to be biased due to researchers’ assumptions about what constitutes an obesity-related cause of death. In light of the continuing obesity epidemic in the United States – with some projections estimating that half of the population will have obesity by the end of the decade (60) – it is critical to have consensus on the definition of obesity-related mortality to produce consistent estimates of its consequences for mortality.

## Data Availability

The code, and all harmonized input and output data pertaining to our analysis, is hosted on GitHub https://github.com/OxfordDemSci/ex_USA.

## Declarations

### Ethics approval and consent to participate

Not applicable.

### Consent for publication

Not applicable.

### Availability of data and materials

The datasets analyzed in this current study are available at NCHS, (https://www.cdc.gov/nchs/data_access/vitalstatsonline.htm) and SEER, (https://seer.cancer.gov/data-software/uspopulations.html).

### Authors contributions

Conceptualization: AMT

Data curation: AMT

Formal analysis: AMT, JMA

Methodology: AMT, JMA, JBD

Software: AMT, JMA

Visualization: AMT, JMA, IG, JBD

Writing – original draft: AMT, IG

Writing – review & editing: AMT, JMA, IG, JBD

## Acknowledgements

We thank the Health Inequality Working Group at the Leverhulme Centre for Demographic Science for their insightful comments on the manuscript.

