## Appendix for "Coding of Obesity-related Mortality Impacts Estimates of Obesity on U.S. Life Expectancy"

**Online Appendix**

I. Appendix A: ICD-10 Codes used for each Coding Scheme
II. Appendix B: ICD-10 Codes used for Other Causes of Death
III. Appendix C. Age Pattern of Obesity-related Mortality across Schemas, 2010-2020

**Appendix A:** ICD-10 Codes used for each Coding Scheme

**Acosta et al.**

| Diabetes | E10-14 |
| --- | --- |
| Obesity | E66 |
| Obesity-related Cancers (esophagus, stomach, colorectal, liver, gallbladder, pancreas) | C15-C16, C18-C25 |

**Adair and Lopez**

| Diabetes | E10-E14 |
| --- | --- |
| Hypertension | I10-I13 |
| Obesity | E65-E66 |
| Chronic Kidney Disease | N18 |
| Lipidemias | E78 |

**Masters et al.**

| Diabetes | E10-E14 |
| --- | --- |
| Heart Disease | I00-I09, I11,I13,I20-I51 |
| Hypertension | I10, I12, I15 |
| Obesity | E65-E67 |

**Global Burden of Disease**

| Neoplasms |  |
| --- | --- |
| Esophageal cancer | C15-C15.9, D00.1, D13.0 |
| Liver cancer | C22-C22.9, D13.4 |
| Breast cancer | C50-C50.929, D05-D05.92,D24-D24.9,D48.6-D48.62,D49.3 |
| Uterine cancer | C54-C54.9,D25-D25.9,N84.0,N87-N87.9 |
| Colorectal cancer | C18-C20.0,C20.9-C21.8,D01.0-D01.3,D12-D12.9,D37.3- D37.5,K62.0,K62.1,K63.5 |
| Gallbladder cancer | C20.8,C23-C24.9,D13.5 |
| Pancreatic cancer | C25-C25.9,D13.6,D13.7 |
| Ovarian cancer | C56-C56.9,D27-D27.9,D39.1-D39.12 |
| Kidney cancer | C64-C65.9,D30.0-D30.12,D41.0-D41.12 |
| Thyroid Cancer | C73-C73.9,D09.3-D09.8,D34-D34.9,D44.0 |
| Leukemia | C91-C95.92 |
| Cardiovascular diseases | G45-G46.8,I01-I01.9,I02.0,I05-I09.9,I11-I11.9,I20-I25.9,I28- I28.8,I30-I31.1,I31.8,I31.9,I33-I42.9,I47-I48.92,I51.0-I51.6,I60- I61.9,I62.0-I62.03,I63-I63.9,I65-I66.9,I67.0-I67.3,I67.5- I67.7,I69.0-I69.198,I69.20-I69.398,I70.2-I70.8,I71-I78.9,I80- I83.93,I86-I89.9,I91.9 |
| Ischemic heart disease | I20-I25.9 |
| Cerebrovascular disease | G45-G46.8,I60-I61.9,I62.0-I62.03,I63-I63.9,I65-I66.9,I67.0- I67.3,I67.5-I67.7,I69.0-I69.198,I69.20-I69.398 |
| Ischemic stroke | G45-G46.8,I63-I63.9,I65-I66.9,I67.2,I67.3,I67.5,I67.6,I69.3- I69.398 |
| Hemorrhagic stroke | I60-I61.9,I62.0-I62.03,I67.0,I67.1,I67.7,I69.0-I69.198,I69.20- I69.298 |
| Hypertensive heart disease | I11-I11.9 |
| Cardiomyopathy | I40-I42.9,I51.4-I51.6 |
| Atrial fibrillation | I48-I48.92 |
| Peripheral vascular | I70.2-I70.799,I73-I73.9 |
| Endocarditis | I33-I33.9,I39-I39.9 |
| Other cardiovascular | I28-I28.8,I30-I31.1,I31.8,I31.9,I34-I38.9,I47-I47.9,I51.0- I51.3,I70.8,I72-I72.9,I74-I78.9,I80-I83.93,I86-I89.9,I91.9 |
| Diabetes/urog/blood/endo | B37.3-B37.49,B37.9,D52.1,D55-D58.9,D59.0- D59.3,D59.5,D59.6,D60-D61.9,D64.0,D64.4,D66-D69.8,D70- D75.89,D76-D78.89,D80-D83.9,D84.0-D84.8,D86.3- D86.87,D89-D89.3,E03-E07.1,E09-E16.9,E20-E34.8,E36- E36.8,E65-E68,E70-E85.29,E87.71,E88-E89.9,G21.1- G21.19,G24.0-G24.09,G25.1,G25.4,G25.6- G25.79,G72.0,G93.7,G97-G97.9,I12-I13.9,I95.2,I95.3,I97- I98.2,I98.9,J70.2-J70.5,J95-J95.9,K43-K43.9,K62.7,K91- K92,K94-K95.89,M87.1-M87.19,N00-N08.8,N10-N12.9,N14- N16.8,N18-N18.9,N20-N23.0,N25-N32.0,N32.3,N32.4,N34- N34.3,N36-N36.9,N39-N39.2,N41-N41.9,N44-N45.9,N49- N51.8,N65-N65.1,N72,N72.0,N75-N77.8,N80-N81.9,N83- N83.9,N99-N99.9,P03.2-P03.5,P70.0- |
| Diabetes | E10-E10.11,E10.3-E11.1,E11.3-E12.1,E12.3-E13.11,E13.3- E14.1,E14.3-E14.9,P70.0-P70.2,R73-R73.9 |
| Chronic kidney disease | E10.2-E10.29,E11.2-E11.29,E12.2,E13.2-E13.29,E14.2,I12- I13.9,N02-N08.8,N15.0,N18-N18.9 |
| Diabetes CKD | E10.2-E10.29,E11.2-E11.29,E12.2,E13.2-E13.29,E14.2 |
| Hypertensive CKD | I12-I13.9 |
| Glomerulonephritis CKD | N03-N06.9 |
| Other CKD | N02-N02.9,N07-N08.8,N15.0 |
| Musculoskeletal disorders | I27.1,L93-L93.2,M00-M03.0,M03.2,M03.6,M05- M09.0,M09.2,M09.8,M30-M32.9,M34-M36.8,M40- M43.19,M65-M65.08,M71.0-M71.19,M86-M87.09,M88- M89.09,M89.5-M89.59,M89.7-M89.9 |

**Appendix B:** ICD-10 Codes used for Other Causes of Death

| **Cause of Death** | **ICD-10 Codes** |
| --- | --- |
| Despair | F10-F16, F19, K70, K73-K74, U03, X40-X45, X64-X85, Y10-Y15, Y87 |
| Cancer | C00-C99 |
| Respiratory Illness | J40-J46 |
| Infectious and Parasitic Diseases | A00-A09, A16-A44, A48-A99, B00-B09, B15-B99, D86.9, G02, G14, H32, I32, I39 J17, K90.8, L44.4, L94.6, M02.3, M35.2, M66, N34.1, R11.1 |
| Accidents | V00-V99, W00-W99, X00-X59, Y85-Y86 (excluding, despair causes) |
| Residual | All else |

**Appendix C.** Age Pattern of Obesity-related Mortality across Schemas, 2010-2020

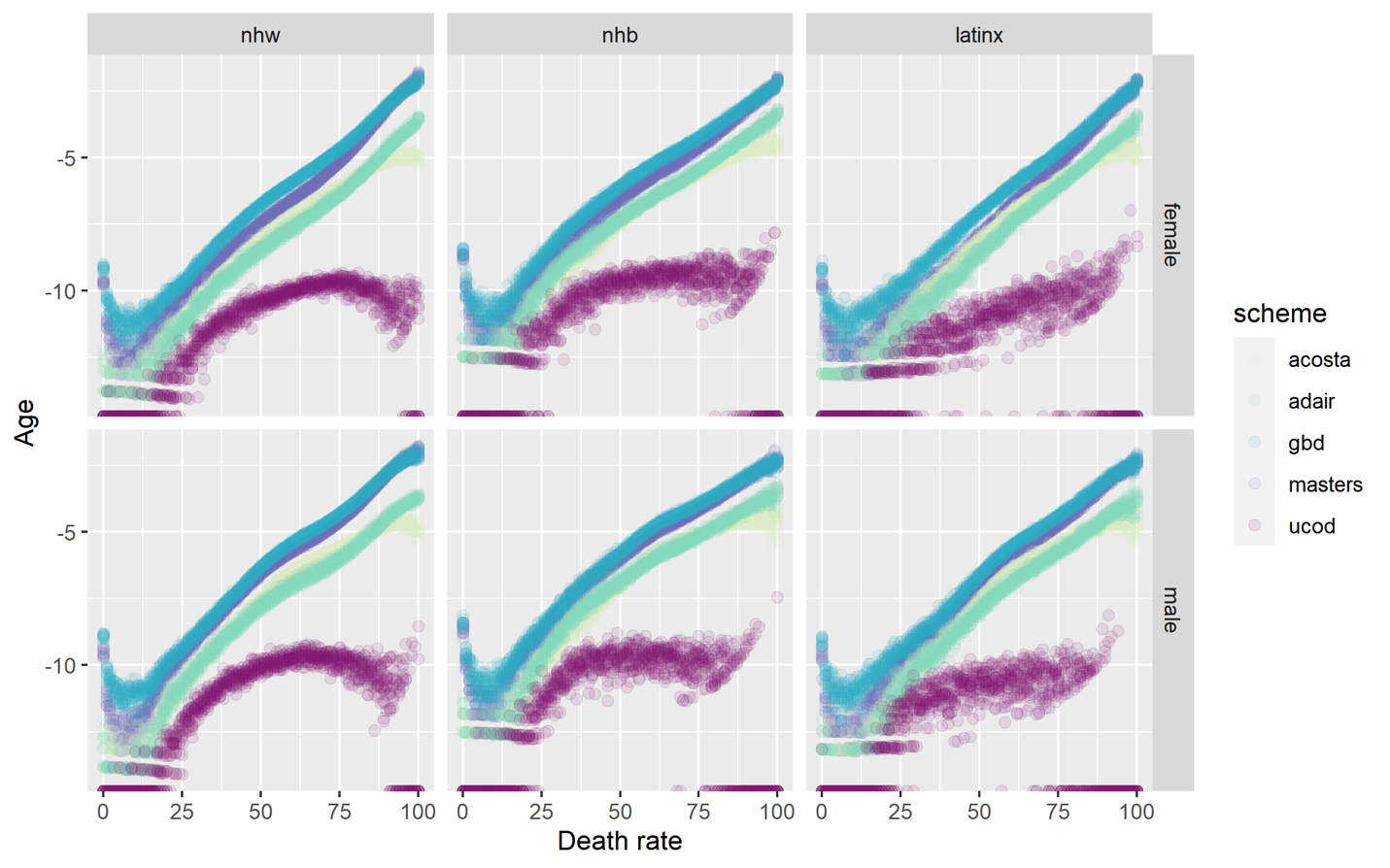
